# Can computer simulation support strategic service planning? Modelling a large integrated mental health system on recovery from COVID-19

**DOI:** 10.1101/2023.07.19.23292289

**Authors:** Livia Pierotti, Jennifer Cooper, Charlotte James, Kenah Cassels, Emma Gara, Rachel Denholm, Richard Wood

## Abstract

**Background:** COVID-19 has had a significant impact on people’s mental health and mental health services. During the first year of the pandemic, existing demand was not fully met while new demand was generated, resulting in large numbers of people requiring support. To support mental health services to recover without being overwhelmed, it was important to know where services will experience increased pressure, and what strategies could be implemented to mitigate this.

**Methods:** We implemented a computer simulation model of patient flow through an integrated mental health service in Southwest England covering General Practice (GP), community-based ‘talking therapies’ (IAPT), acute hospital care, and specialist care settings. The model was calibrated on data from 1 April 2019 to 1 April 2021. Model parameters included patient demand, service-level length of stay, and probabilities of transitioning to other care settings. We used the model to compare ‘do nothing’ (baseline) scenarios to ‘what if’ (mitigation) scenarios, including increasing capacity and reducing length of stay, for two future demand trajectories from 1 April 2021 onwards.

**Results:** The results from the simulation model suggest that, without mitigation, the impact of COVID-19 will be an increase in pressure on GP and specialist community based services by 50% and 50-100% respectively. Simulating the impact of possible mitigation strategies, results show that increasing capacity in lower-acuity services, such as GP, results in demand being shifted to other parts of the mental health system while decreasing length of stay in higher acuity services is insufficient to mitigate the impact of increased demand.

**Conclusion:** In capturing the interrelation of patient flow related dynamics between various mental health care settings, we demonstrate the value of computer simulation for assessing the impact of interventions on system flow.

## Background

The COVID-19 pandemic has had a significant impact on people’s mental health and mental health services. During the pandemic, difficulties in accessing services have suppressed existing demand while new demand has been generated by the social and financial consequences of lockdown, bereavement, virus anxiety, and trauma in healthcare workers (Jia et al., 2020; Mazza et al., 2020; Shevlin et al. 2020). Consequently, as mental health services re-open, a surge in demand is expected (Xiang et al., 2020). For mental health services to recover from the pandemic and meet demand, knowing which components of a service will experience increased pressure is essential, alongside whether changes in service design could mitigate such effects.

Computer simulation, a digital model that replicates real-life processes, has a proven track record in informing and improving the management of health services (Salleh et al, 2017; Mohiuddin et al, 2020; Vázquez-Serrano et al,2021). Simulations can be used to understand relationships, feedback pathways and processes across multi-organisation systems, and assess how these would behave if a change occurred. As they are both safer and cheaper than conducting experiments in a real-world setting, they are useful for testing the potential impact of changes in service design.

Simulation modelling has been under-used in mental health service planning and development, compared to other clinical disease pathways (Long, 2017). While existing studies have implemented system dynamic modelling for improving mental health services during the pandemic (Katikireddi, 2022; Currie et al, 2020), simulation has only previously been applied to decision-making in treatment evaluation, cost-effectiveness analysis and epidemiological studies (Long, 2017).

In this study, we use a Discrete Time Simulation (DTS) model to evaluate the impact of the COVID-19 pandemic on mental health services. Using the model, we assess where the pandemic-related surge in demand will lead to increased pressure, and the effect of possible mitigation strategies to reduce this pressure. Our study demonstrates how simulation modelling can be used to inform decisions regarding changes to capacity and structure of mental health pathways, to best meet the needs of patients on recovery from the pandemic and beyond.

To address these questions, we first described the flow of patients across multiple mental health services in the system by developing a schematic representation of the mental health pathways using linked electronic patient-level data up to 1 April 2021. The pathway maps resulting from this exercise in ‘process mining’ were used to configure the structure of the DTS, in terms of the mental health services covered – GP, Improving Access to Psychological Therapy (IAPT) care, acute hospital, and specialist care settings – and the various parameters relating to patient flow between these services – arrival rates, lengths of stay, and transition probabilities. The model also captures the effect of escalating need should demand not be met in a timely manner. With the model calibrated on data to 1 April 2021, this study then explores the potential effects of two different scenarios relating to future mental health demand on recovery from the Covid-19 pandemic; first on a ‘do nothing’ (baseline) basis, and then through considering service-level mitigatory measures through a ‘what if’ (intervention) analysis. In addition, baseline and intervention scenarios can be compared in terms of whether needs are adequately met for each patient, which can be evaluated by looking at patients exiting the pathway without having received treatment.

## Methods

### Study Setting

Bristol, North Somerset, and South Gloucestershire (BNSSG) Integrated Care System (ICS) is a healthcare system in southwest England, with a population of approximately one million residents. As with other NHS systems, BNSSG ICS is a network of healthcare providers covering primary, secondary, mental health, community, and social care. The healthcare system serves a mixture of metropolitan areas and rural and coastal locations. The large metropolitan area of Bristol contains a higher proportion of younger individuals and is culturally and ethnically diverse. Rural and coastal areas contain a greater proportion of older individuals and pockets of severe deprivation (Ministry of Housing Communities & Local Government, 2019).

### Population

The study population included all patients aged 18 years and over referred to mental health services in BNSSG between 1 April 2019 to 1 April 2021. Patients were excluded if they were in community mental health services for children.

### Data

Analysis was conducted using the BNSSG System Wide Dataset (SWD) from 1 April 2019 to 1 April 2021. The SWD provides patient-level linkable primary, secondary and community health data for the BNSSG population (BNSSG Healthier Together, 2021). Primary Care data is obtained via a bespoke extract from general practitioners, collated by OneCare, which is a General Practice (GP) federation operating in BNSSG. Sourced from Egton Medical Information Systems (EMIS) GP administration systems, the extract contains data on GP attendances and prescriptions. Secondary Uses Service (SUS) contains information on all NHS acute trust outpatient consultations, inpatient admissions, and emergency department attendances, with detailed data on date of attendance, ward specialty and clinical indications. The Community Services Data Set (CSDS), maintained by NHS Digital, includes intermediate care admissions and patient visits to and from community service teams. Mental health data, covering consultations and admitted stays, is available from the Mental Health Services Data Set (MHSDS) also maintained by NHS Digital. A full specification of the System Wide Dataset is publicly available (BNSSG Healthier Together, 2021) including the data dictionary. To mitigate any risks associated with the holding of patient identifiable data, all records are pseudonymised by the regional Commissioning Support Unit before being added to the SWD. The dataset also contains no patient names or full addresses.

The SWD contains two data tables: attributes and activities, linkable using the pseudonymised patient identifier. The attribute table is a monthly data flow of social demographic factors (i.e., sex, age, LSOA, ethnicity) and clinical factors from patients registered to participating GPs within BNSSG. The activity table contains information on date of medical appointment, prescription and the specific healthcare service that the patient had appointment for.

The SWD was linked to IAPT care data. The IAPT programme is a large-scale initiative that aims to greatly increase the availability of National Institute for Health and Care Excellence (NICE) (NICE, 2011) recommended psychological treatment for depression and anxiety disorders within the NHS. It offers a range of talking therapies in addition to those that the NHS can offer. This includes interpersonal therapy, couples therapy, and counselling for depression. Through IAPT services, approximately one million individuals per year start treatment (NHS Digital, 2020). A full description of measures and conditions treated under IAPT services can be found via the IAPT manual (National Collaborating Centre for Mental Health, 2019).

### Derived variables

#### Clinical severity

Using the attribute table within the SWD, we define mental health severity using 3 categories: severe, moderate and mild. A patient’s mental health severity is categorised as severe if they are suffering from a chronic mental health condition, such as depression, post-traumatic stress disorder or an eating disorder. A patient without a diagnosis of a mental health condition, but living with an associated condition, such as drug or alcohol dependency, autism, or ADHD, is categorised as moderate. Patients without any specific mental health or associated diagnoses are categorised as mild.

### Level of care

To enable mental health clinical pathways to be mapped across the system, services were grouped into six levels according to clinical need (Figure 1), defined by clinical, managerial, and analytical stakeholders from across the BNSSG mental health system. Level 1 represents the lowest level of care required and 6 the highest. Of the six levels of care identified, only services in levels 2-5 were included in the study. Level 1 services (community support) were excluded as they cover a very broad array of services. It was not possible to recover reliable data for level 6 services.

**Figure 1:**
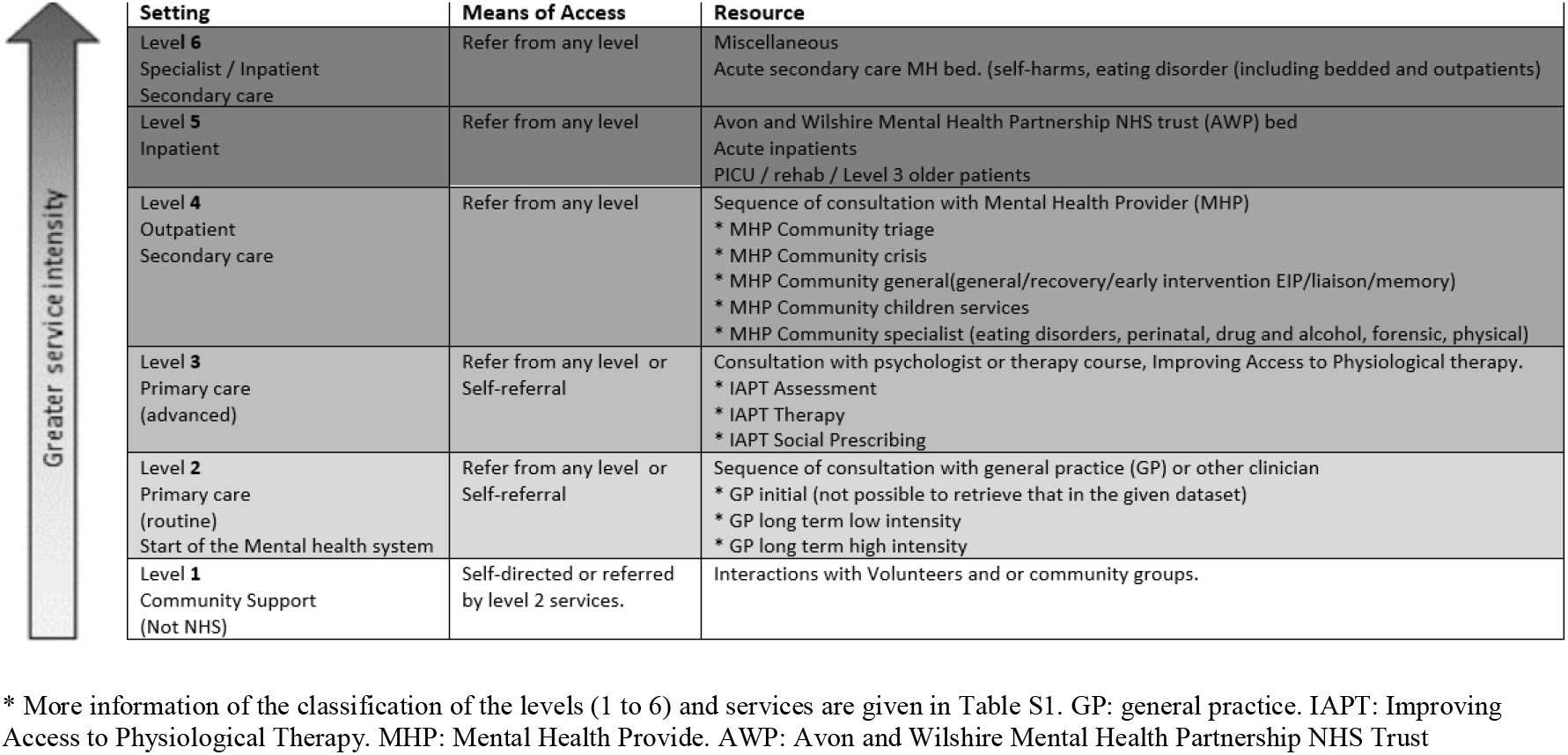
Description of six ‘levels of need’ for mental health services in the healthcare system studied, detailing the setting, means of access, and type of resource. The colours represent the level of intensity of care services, where the lighter colours represent the least intensity.

### Process mining

Patients experience mental health care pathways as causally linked sequences of activities. However, in the data, these activities are recorded separately as discrete events. To reconstruct patient pathways from the SWD and IAPT data, we implemented process mining, a technique commonly used for extracting clinical pathways from administrative data (Rojas et al, 2016).

The mental health pathway in BNSSG, obtained through process mining, is displayed as a network map in Figure 2. Here, nodes represent services within the pathway and directed edges represent possible routes a patient can take between services. Using the reconstructed pathways, we calculated arrival rates, lengths of stay, and transition probabilities between levels of care and services to be used as parameters of the DTS. We also calculated the waiting time for each service and the ’reneging’ rate – i.e., the rate at which patients leave the waiting list for a specific service without treatment and are transferred elsewhere – as additional DTS parameters.

**Figure 2:**
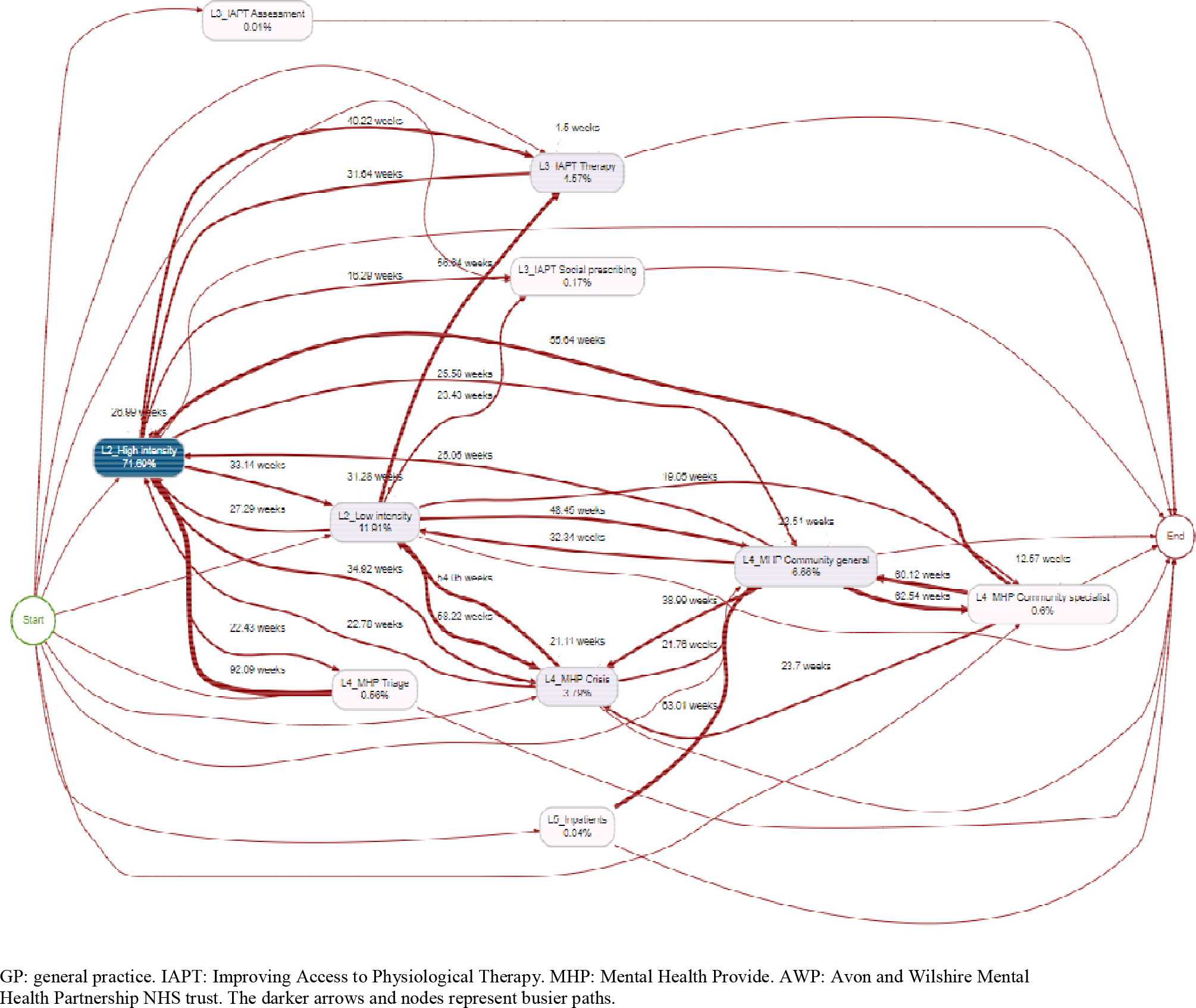
Figure and online schematic representation (NHS BNSSG Analytics, 2022) of a sample of 10,000 entries of the mental health services to be modelled. Service nodes were aligned to the six considered levels of need as detailed in Figure 1. GP: General Practice; IAPT: Improving Access to Psychological Therapies; MHP: Mental Health Provider.

### Computer simulation

A DTS model, developed by Murch et al (2021), was used to simulate patient flow across the reconstructed mental health pathway. The DTS models a patient’s pathway through the mental health service as a series of events. Both the time between two events, and the next event in a patient’s pathway, are determined by sampling from probability distributions. The probability distributions are parameterised by the arrival rates and lengths of stay for each service, and transition probabilities between services, obtained from the reconstructed pathway. For a technical description of the DTS methodology see Murch et al (2021).

Events simulated in the DTS include: a patient previously not known to mental health services presenting with a mental health condition; a patient moving from one service to another; a patient queuing for capacity in a service. Patients enter the model when there is available capacity in a service. The time a patient remains in a service is sampled from the length of stay distribution for that service. After a patient’s length of stay has elapsed, they move to a different service. The service a patient moves to is determined using the transition probabilities between the service they are in and all other services. Patients leave the model if they are either successfully treated or are referred to another service for ongoing care. To capture patient deconditioning associated with long waits incurred during the pandemic, after a specified amount of time waiting for a service, patients can ‘renege’ from the queue they are in and join the pathway of a higher-level service or discharge themselves from the pathway without treatment.

Using the DTS, patient events were simulated for each day within the post-lockdown study period (Figure 3). For each simulated day, the number of patients in each service, the number of patients waiting for each service, and the number of patients that reneged were recorded. This resulted in a time series for each of these measures over the study period.

**Figure 3:**
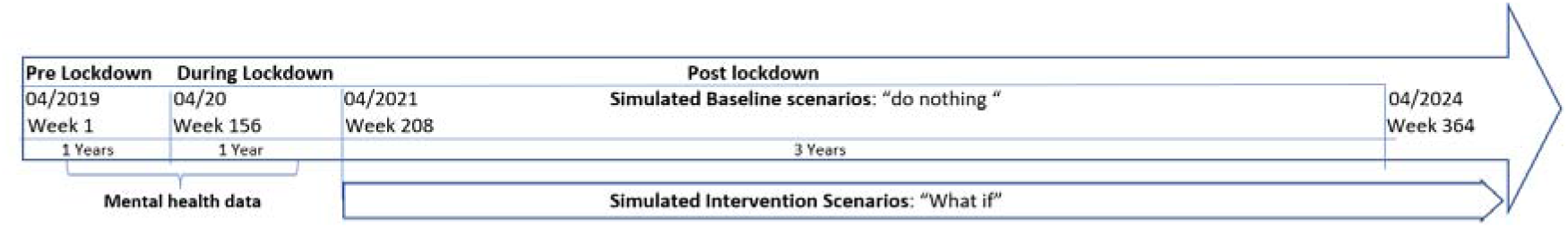
Scenario Timeline. The study used data for the pre and during lockdown periods (from 1 April 2019 to 1 April 2021, representing week 1 to week 207 of the study period) to obtain model parameters, and simulated baseline and intervention scenarios for the post lockdown period (from 1 April 2021 to 1 April 2024, from week 208 to week 364 of the study period assumed for this study).

As daily arrivals at each service, lengths of stay, and service transitions are randomly sampled from distributions, one simulation of the study period represents only one way in which events could pan out. To capture the range of possible outcomes over the study period, 35 replications of the simulation were performed, each using a different random seed. Results from each simulation were averaged over all replications.

### Scenarios

The DTS was used to evaluate scenarios to mitigate waiting time and capacity at different mental health services after lockdown. Four scenarios were modelled: two baseline and two interventions. Baseline scenarios represent hypothesised changes to patient flow post-lockdown (from April 2021), as described in the literature (Tojesen, 2020; Hood et al, 2020). The demand profiles (i.e., external arrival rates) in the DTS were adjusted according to each baseline scenario to model the different problem(s) that mental health services might encounter, such as where large queues may form, as they recover from the pandemic. For each baseline scenario, a plausible intervention scenario was simulated to estimate the impact possible measures to mitigate pressure could have (Table 1). The DTS was used to simulate demand trajectories from the 1 April 2021 to April 2024 (Figure 3) for each scenario.

**Table 1:**
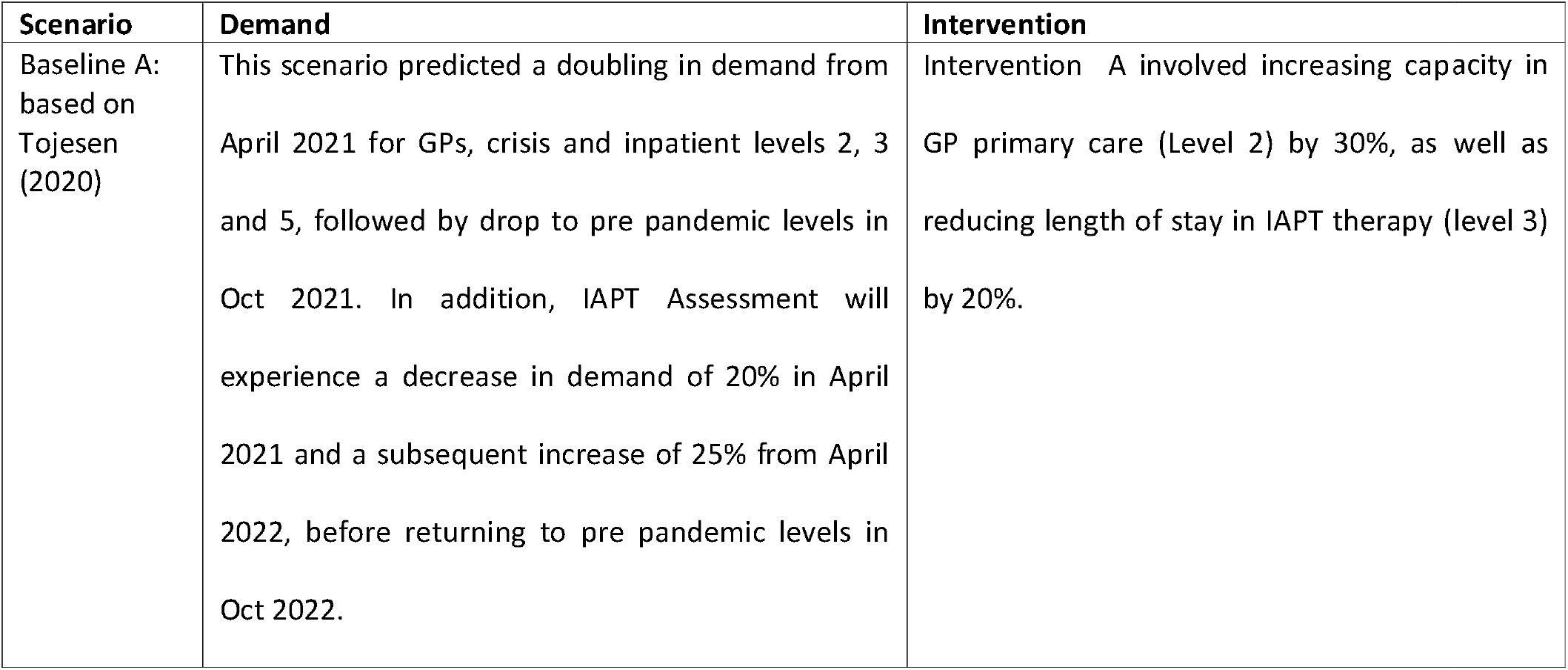
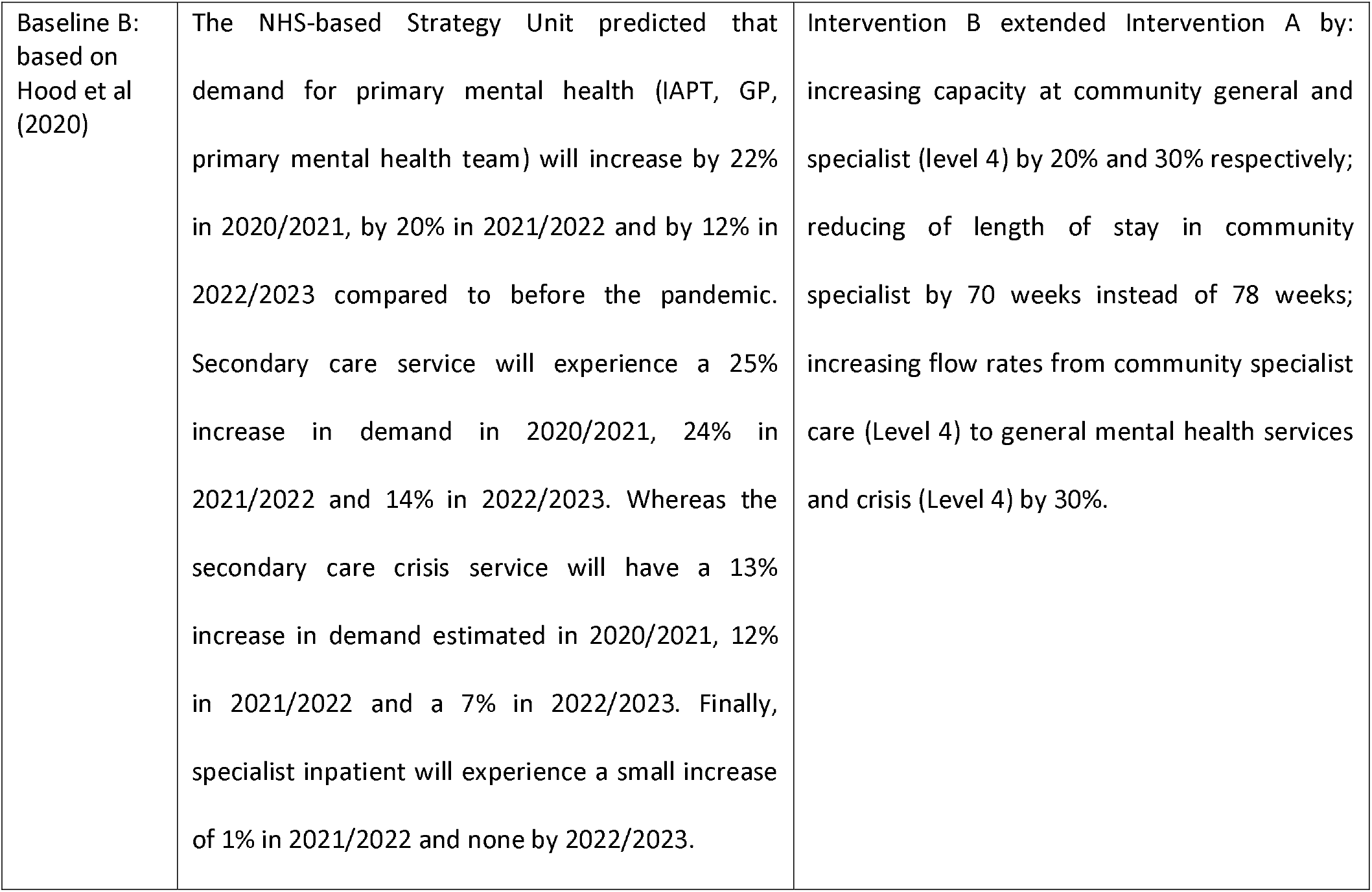
The four simulation scenarios considered in this study (two baseline scenarios and two intervention scenarios). Baseline scenarios were obtained from literature. The two intervention scenarios were related to the results of the two simulated baseline scenarios.

The DTS was used to simulate baseline scenarios and assess the impact of increased demand on patient waiting times, service occupancy, and reneging rate. Estimates of demand in each service (mean and 95% confidence intervals (CI)) were obtained from 35 replications of the DTS. Estimated demand profiles were scaled by true demand in each service on 1 April 2021, allowing future demand to be assessed relative to this timepoint.

Intervention scenarios (Intervention A and B, Table 1) were used to investigate the impact of increasing capacity, reducing length of stay or re-routing patients to different services, on service occupancy, waiting times and reneging rate.

## Results

### Data

There were 289,666 attendances by 188,682 patients recorded in the mental health pathway between 1 April 2019 and 1 April 2021 (Table 2). Of the attendances, 254,208, (88%) were in primary care level 2 and level 3, 33,830, (12%) were in secondary care outpatient level 4 and 1% were in AWP inpatients level 5. 100,915 (35%) of the patients within the pathway were referred during April 2020 to April 2021. The cohort was predominately female (61%) and white (85%) and aged 40–59-year-old (30%). Those who needed the highest level of service (level 4 and 5), were from the most deprived groups. Almost 90% of the patients waiting at level 5 inpatients have experienced at least one moderate mental health condition and 26% of those had experience at least one severe mental health condition.

**Table 2.** Demographic of study patients by level of need.

|  | L2 GP<br>High<br>Intensity | L2 GP<br>Low<br>Intensity | L3 IAPT<br>Assessment | L3 IAPT<br>Social<br>prescribing | L3 IAPT<br>Therapy | L4 MHP<br>General | L4 MHP<br>Specialist | L4 MHP<br>Crisis | L4 MHP<br>Triage | L5 AWP<br>Inpatients |
| --- | --- | --- | --- | --- | --- | --- | --- | --- | --- | --- |
| N= | 96169 | 123146 | 410 | 2928 | 31555 | 9720 | 3219 | 10049 | 10842 | 1628 |
| Activity Volume in pre-pandemic | 61729 | 87385 | 404 | 2016 | 16180 | 5303 | 2025 | 6119 | 6557 | 1033 |
| Activity volume during lockdown | 34440 | 35761 | 6 | 912 | 15375 | 4417 | 1194 | 3930 | 4285 | 595 |
| Sex |  |  |  |  |  |  |  |  |  |  |
| Female | 59488 (61.9) | 73256 (59.5) | 287 (70.0) | 1980 (67.6) | 21172 (67.1) | 5481 (56.4) | 1937 (60.2) | 5489 (54.6) | 6262 (57.8) | 770 (47.4) |
| Male | 36671 (38.1) | 49880 (40.5) | 123 (30.0) | 948 (32.4) | 10379 (32.9) | 4237 (43.6) | 1280 (39.8) | 4558 (45.4) | 4579 (42.2) | 856 (52.6) |
| Age |  |  |  |  |  |  |  |  |  |  |
| 18-29 | 22617 (23.5) | 31715 (25.8) | 107 (26.1) | 915 (31.2) | 11748 (37.2) | 2052 (21.1) | 1485 (46.1) | 2688 (26.7) | 3528 (32.5) | 311 (19.1) |
| 30-39 | 19168 (19.9) | 25432 (20.7) | 96 (23.4) | 777 (26.5) | 8728 (27.7) | 843 (8.7) | 1054 (32.7) | 2009 (20.0) | 2356 (21.7) | 342 (21.0) |
| 40-59 | 29457 (30.6) | 38419 (31.2) | 147 (35.9) | 937 (32.0) | 8344 (26.4) | 1239 (12.7) | 605 (18.8) | 2624 (26.1) | 2970 (27.4) | 552 (33.9) |
| 60-79 | 17925 (18.6) | 20876 (17.0) | 56 (13.7) | 280 (9.6) | 2479 (7.9) | 2062 (21.2) | 74 (2.3) | 1501 (14.9) | 1224 (11.3) | 320 (19.7) |
| 80-90+ | 7002 (7.3) | 6704 (5.4) | 4 (1.0) | 19 (0.6) | 256 (0.8) | 3524 (36.3) | 1 (0.0) | 1227 (12.2) | 764 (7.0) | 103 (6.3) |
| Ethnicity |  |  |  |  |  |  |  |  |  |  |
| Asian / Asian British | 2330 (2.8) | 2910 (2.7) | 1 (0.3) | 71 (2.8) | 731 (2.7) | 144 (1.8) | 56 (2.1) | 213 (2.5) | 197 (2.1) | 42 (3.0) |
| Black/African/Caribbean/Black British | 2031 (2.5) | 2727 (2.6) | 2 (0.6) | 53 (2.1) | 644 (2.4) | 200 (2.4) | 45 (1.7) | 285 (3.3) | 204 (2.2) | 96 (6.9) |
| Mixed / Multiple ethnic groups | 5066 (6.1) | 7169 (6.8) | 18 (5.1) | 144 (5.7) | 2084 (7.6) | 423 (5.2) | 224 (8.4) | 612 (7.2) | 607 (6.6) | 92 (6.6) |
| Other / Unknow | 2404 (2.9) | 3019 (2.9) | 3 (0.8) | 61 (2.4) | 804 (2.9) | 261 (3.2) | 62 (2.3) | 303 (3.5) | 308 (3.3) | 41 (2.9) |
| White | 70715 (85.7) | 90025 (85.0) | 329 (93.2) | 2204 (87.0) | 22997 (84.4) | 7151 (87.4) | 2276 (85.5) | 7131 (83.5) | 7894 (85.7) | 1124 (80.6) |
| SES<sup>1</sup> |  |  |  |  |  |  |  |  |  |  |
| 1 (most deprived) | 19928 (20.7) | 26645 (21.7) | 89 (21.7) | 610 (20.8) | 6605 (21.0) | 2185 (22.5) | 943 (29.3) | 2765 (27.6) | 2617 (24.2) | 532 (32.7) |
| 2 | 17457 (18.2) | 23606 (19.2) | 64 (15.6) | 591 (20.2) | 6450 (20.5) | 1884 (19.4) | 653 (20.3) | 2096 (20.9) | 2201 (20.3) | 385 (23.7) |
| 3 | 16379 (17.0) | 20716 (16.8) | 64 (15.6) | 487 (16.6) | 5524 (17.5) | 1600 (16.5) | 536 (16.7) | 1741 (17.4) | 1963 (18.1) | 228 (14.0) |
| 4 | 20856 (21.7) | 25779 (21.0) | 111 (27.1) | 637 (21.8) | 6709 (21.3) | 1996 (20.6) | 596 (18.5) | 1871 (18.7) | 2050 (18.9) | 263 (16.2) |
| 5 (least deprived) | 21478 (22.4) | 26279 (21.4) | 82 (20.0) | 603 (20.6) | 6235 (19.8) | 2044 (21.1) | 486 (15.1) | 1558 (15.5) | 1995 (18.4) | 218 (13.4) |
| mental health condition |  |  |  |  |  |  |  |  |  |  |
| Severe<sup>2</sup> | 14139 (14.7) | 17609 (14.3) | 121 (29.5) | 409 (14.0) | 4328 (13.7) | 1922 (19.8) | 1017 (31.6) | 2364 (23.5) | 2669 (24.6) | 426 (26.2) |
| Moderate<sup>3</sup> | 65384 (68.0) | 88657 (72.0) | 357 (87.1) | 2272 (77.6) | 24378 (77.3) | 5753 (59.2) | 2516 (78.2) | 8730 (86.9) | 9607 (88.6) | 1459 (89.6) |
| Mild<sup>4</sup> | 89 (0.1) | 122 (0.1) | 1 (0.2) | 4 (0.1) | 35 (0.1) | 33 (0.3) | 23 (0.7) | 50 (0.5) | 50 (0.5) | 9 (0.6) |

<sup>1</sup>SES: socioeconomic status. <sup>2</sup>Severe condition (Associated condition, i.e., Autism, ADHD, addiction) <sup>3</sup>Moderate condition (i.e., depression, anxiety, eating disorder). <sup>4</sup>Mild condition (Social Factors, i.e., homeless). GP: general practice. IAPT: Improving Access to Physiological Therapy. MHP: Mental Health Provide. AWP: Avon and Wilshire Mental Health Partnership NHS trust.

### Baseline Simulation Modelling

Results from using the DTS to model waiting list size, service occupancy and reneging rates under Baseline Scenarios A and B are displayed in Figure 4. Simulation results suggest the waiting list size (Figure 4, top row), when compared to pre-pandemic size, will increase in level 2 services (GP) by up to 50% for both baseline scenarios. Under Scenario B, results suggest there will also be moderate increases in waiting list size for level 3 services (IAPT) and increases of 50-150% in community based (level 4) mental health services. As a result, the occupancy of these specific services tends to increase (Figure 4, second row), creating blockages because the services do not have the capacity to handle the sharp increase in demand post-lockdown. Due to increased waiting times and high service occupancy in community services, results suggest an increased tendency to renege from these services under both scenarios (Figure 4, third row). Both baseline scenarios showed an increase in reneging to GP services, whereas only Scenario B showed an increase in reneging to MHP crisis (Figure 4, bottom row).

**Figure. 4:**
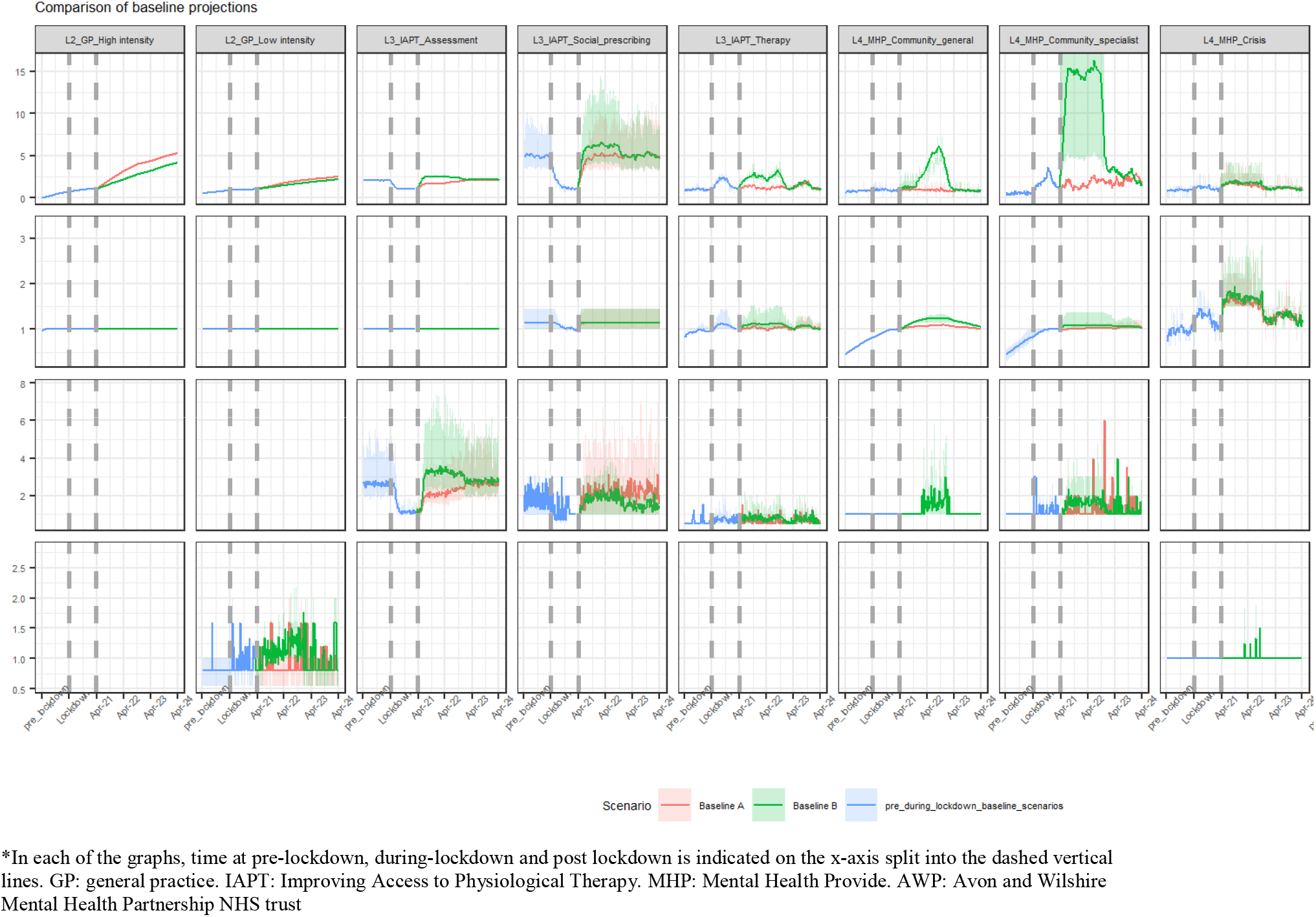
Summary of the simulation results of the queue size, occupancy, and renege output measures at each service (per week) for Baseline Scenarios A (red) and B (green), modelled pre-, during, and post-lockdown. Solid lines represent mean demand profiles obtained from 35 replications of the DTS, relative to demand at the end of the lockdown period, and the shaded area represents a 95% confidence interval in scaled demand. Results were omitted for services which display negligible change over the course of these three periods.

### Intervention Simulation Modelling

Table 3 shows the results from using the DTS to simulate two intervention scenarios for mitigating increased pressure following lockdown. To estimate the effectiveness of each intervention, we report the percentage change in the mean and maximum values of waiting list size, service occupancy and reneging rates compared to the baseline scenarios.

**Table 3:** Summary of the impact of Interventions A and B on mental health system performance from 1 April 2021. Results represent the maximum and mean percentage change between baseline and intervention scenarios.

| Service node | Measure<br>Change in | Intervention A |  |  |  | Intervention B |  |  |  |
| --- | --- | --- | --- | --- | --- | --- | --- | --- | --- |
|  |  | Waiting<br>List<br>(%) | Service<br>Occupancy<br>(%) | renege<br>from<br>(%) | renege<br>to<br>(%) | Waiting<br>List<br>(%) | Service<br>Occupancy<br>(%) | renege<br>from<br>(%) | renege<br>to<br>(%) |
| L2 GP High intensity | max | -5 | 6 | - | - | -4 | 0 | - | - |
|  | mean | -7 | 1 | - | - | -5 | 0 | - | - |
| L2 GP Low intensity | max | 0 | 8 | - | 20 | 0 | 1 | - | 0 |
|  | mean | 0 | 1 | - | 3 | 0 | 0 | - | -10 |
| L3 IAPT Assessment | max | 0 | 0 | -2 | - | 0 | 0 | 0 | - |
|  | mean | 0 | 0 | 0 | - | 0 | 0 | 0 | - |
| L3 IAPT Social prescribing | max | 0 | 0 | -4 | - | 0 | 0 | 14 | - |
|  | mean | 0 | 0 | -2 | - | -1 | 0 | 5 | - |
| L3 IAPT Therapy | max | 69 | 2 | 9 | - | 19 | 2 | 20 | - |
|  | mean | 44 | 3 | 17 | - | 17 | 2 | -5 | - |
| L4 MHP general | max | 205 | 16 | - | - | 194 | 4 | 100 | - |
|  | mean | 109 | 16 | - | - | 43 | 3 | 2 | - |
| L4 MHP specialist | max | 29 | 2 | -43 | - | -20 | -1 | -50 | - |
|  | mean | 37 | 2 | 0 | - | -34 | -4 | -14 | - |
| L4 MHP Crisis | max | 136 | 18 | - | 0 | 81 | 18 | - | 0 |
|  | mean | 11 | 7 | - | 0 | 20 | 9 | - | 0 |
| L4 MHP Triage | max | 215 | 63 | - | - | 160 | 40 | - | - |
|  | mean | 17 | 8 | - | - | 11 | 4 | - | - |
| L5 AWP Inpatients | max | -2 | -2 | - | - | 0 | -4 | - | - |
|  | mean | 0 | -3 | - | - | -4 | 3 | - | - |

GP: general practice. IAPT: Improving Access to Physiological Therapy. MHP: Mental Health Provide. AWP: Avon and Wilshire Mental Health Partnership NHS trust

Given the large increase in waiting times for GP (level 2) services post-lockdown under Baseline Scenario A, Intervention A focusses on increasing capacity in GP services to reduce waiting times and decreasing length of stay in IAPT therapy and level 4 services. The purpose of decreasing length of stay in these services is to prevent demand being shifted from level 2 services because of increased capacity. To simulate Intervention A using the DTS, capacity in level 2 services was increased by 30% for the period April 2021 to October 2022 and length of stay in IAPT therapy and level 4 services was decreased by 20% for the period April 2022 to October 2022.

Results from the DTS show that increasing GP capacity would lead to waiting list size for high intensity GP services decreasing by an average of 7% compared to Baseline Scenario A and service occupancy would increase by only 1% (Table 3). However, despite decreasing length of stay in IAPT therapy and level 4 services, simulations show demand would still be shifted: waiting list size would increase by over 10% in all of these higher level services (Table 3, Figure S1). This suggests that reducing length of stay by 20% is insufficient to absorb the extra demand on these services generated by increased capacity in level 2. Results from the DTS show increased capacity in level 2 does, however, have a positive impact on reneging: decreasing waiting times for GP services leads to fewer patients reneging compared to Baseline Scenario A (Table 3).

Intervention B was an extension of Intervention A, to mitigate the impact of both the increased demand due to lockdowns, and the shifts in demand from level 2 to level 4 services following Intervention A. To decrease the waiting times for services that were blocked as a consequence of the mitigations in Intervention A, Intervention B additionally involved: increasing capacity at MHP community general and specialist services by 20 and 30 % respectively ; decreasing length of stay by 10% in the MHP community specialist; increasing patient flow by 30 % from MHP community specialist to MHP general mental health and crisis services. These additional interventions were simulated for the period April 2021 to October 2022.

Using the DTS to simulate Intervention B, results show a decrease in waiting list size, occupancy, and reneging rate for the MHP specialist service compared to Baseline Scenario B (Table 3, Figure S2). Compared to Intervention A, the additional measures are successful in mitigating some of the pressure on the IAPT therapy service, however both waiting times and service occupancy are greater than Baseline Scenario B (Table 3).

## Discussion

Our work demonstrates how simulation modelling may be used to assess the impact of interventions on mental health service pathways. We used process mining to develop a schematic representation of mental health pathways using linked electronic patient-level data up to April 2021. The resulting pathway maps were used to obtain parameters of a DTS model, in terms of mental health services covered (GP, IAPT) care, acute hospital, and specialist care settings) and patient flow between these services (arrival rates, lengths of stay, and transition probabilities). The model also captures the effect of escalating need should demand not be met in a timely manner (reneging). We calibrated the model with data to April 2021, and used it to assess the impact future demand may have on mental health services under different scenarios; first on a ‘do nothing’ basis (Baseline Scenarios A and B), and through considering service-level mitigatory measures through a ‘what if’ analysis (Intervention scenarios A and B).

To model mental health service demand following the pandemic, we used two different baseline scenarios influenced by existing literature (Tojesen, 2020; Hood et al, 2020). This allowed us to forecast demand at each service during a time when the true impact of COVID on mental health services was unknown and derive realistic intervention scenarios to mitigate the impact of this demand. On comparing the intervention scenarios with baseline projections, we found that the intervention scenarios considered in our study are not sufficient for mitigating pressure due to increased demand on mental health services following lockdown. Instead, we found that while pressure may be reduced in one service, the consequence is an increase in pressure in other services. These results suggest that isolated capacity increases in particular parts of a mental health pathway do not necessarily benefit the system: changes in capacity can unblock part of a pathway, allowing unserved demand to flow into other services, increasing utilization and queueing. Our results highlight the need for strategic decision making around changes to service capacity, to ensure that improvements in one service do not have a negative impact on other services and wider system flow.

For both intervention scenarios, we found that increasing capacity in level 2 would reduce waiting list size and service occupancy for these services. As this change to the pathway had a negative impact on higher level services (IAPT Therapy and level 4 services, Table 3), one extension to the interventions considered would be to increase capacity in these services in addition to level 2. However, in the local system the plausibility of this increase is unrealistic: simulations of baseline scenarios suggest a 50-150% increase in demand for level 4 services without any changes to level 2 capacity (Figure 4). Increasing existing capacity in higher level services to meet this increased demand is therefore not a plausible intervention, and as with increases to GP capacity, changes may result in blockages further along the pathway requiring further capacity increases.

A key assumption of the DTS used in this study is that a patient can only be in one service at any given time. While this assumption allows us to model mental health services using DTS, it is also a limitation of the approach: in mental health systems patients can simultaneously be in receipt of care from multiple services at any one time. A further assumption of the model is that patients queuing for a service are seen on a first-come first-served basis. However, when patients are referred to a mental health service, they are assigned a priority which determines the order in which they are seen: patients are not seen in the order in which they are referred (Dehghan et al, 2017). In the future, the DTS framework implemented in this study could be extended to relax these assumptions. Extending the framework in this way would improve the accuracy of simulations and ensure results are representative of the way in which mental health services operate.

In the UK, the use of simulation modelling for mental health service design and delivery is limited by the availability of high-quality data (Jacobs et al, 2019). Data capture and linkage across services means there are often large gaps and inconsistencies in patients’ pathways. As this data was required for validating the model, for our study the help of system stakeholders with expert knowledge of service delivery was necessary for parameterisation of the model. This highlights the need for improvements in data capture and linkage with healthcare systems: incentives to improve data quality in local systems will ensure computational techniques, such as simulation modelling, can be utilised for service improvement.

Our work demonstrates the potential value of computer modelling and simulation for supporting strategic service planning within mental health care. Highlighting the benefits of simulation, the approach contained in this paper may serve as a blueprint for conducting similar modelling exercises in any healthcare systems. With regards to system recovery following the pandemic, we have demonstrated the potential of simulation as an aid to strategic decision making and service planning: modelling realistic scenarios for local healthcare systems can provide useful and actionable insight to clinicians and managers on the ground at a time crucially important for effective future planning.

## Conclusion

In this study, we have demonstrated the value of simulation modelling for assessing the impact of changes in service delivery on mental health pathways. Our results have informed decisions regarding the resourcing and restructuring of capacity and pathways in the local mental health service, to best meet the needs of patients during the pandemic and beyond.

## Data Availability

No data are available. This study used a discrete event simulation tool purpose built for modelling patient pathways. Model code used for this study is freely available at https://github.com/nhs-bnssg-analytics.

https://github.com/nhs-bnssg-analytics/simulation-dts-renege

## List of abbreviations

NHS: National Health Scotland
BNSSG ICS: Bristol, North Somerset and South Gloucestershire healthcare Integrated Care System
SWD: System Wide Dataset
MH: mental health
AWP: Avon and Wilshire Mental Health Partnership NHS trust
MHP: Mental Health Provide
IAPT: Improving Access to Physiological Therapy
GP: general practice
CI: Confidence Intervals
DTS: Discrete time simulation.

## Ethics approval and consent to participate

## Consent for publication

## Availability of data and materials

This study used a discrete event simulation tool purpose built for modelling patient pathways. Model code used for this study is freely available at https://github.com/nhs-bnssg-analytics.

## Authors’ contributions

Concept, methodology and design: Wood, Pierotti, Cooper, Data curation: Cooper, Pierotti

Analysis and interpretation of data: Pierotti, Wood, Cassels, Gara

Writing-original draft preparation: Pierotti Software, Validation: Wood, Pierotti

Writing reviewing and editing: Wood, Denholm, James Funding acquisition: Denholm, Wood, Cooper

Project administration: Cooper, Pierotti Supervision: Wood

## Acknowledgements

This work was supported by the Elizabeth Blackwell Institute, University of Bristol, the Welcome Trust Institutional Strategic Support Fund and the Rosetrees Trust.

## Supplementary tables and figures

**Table S1:**
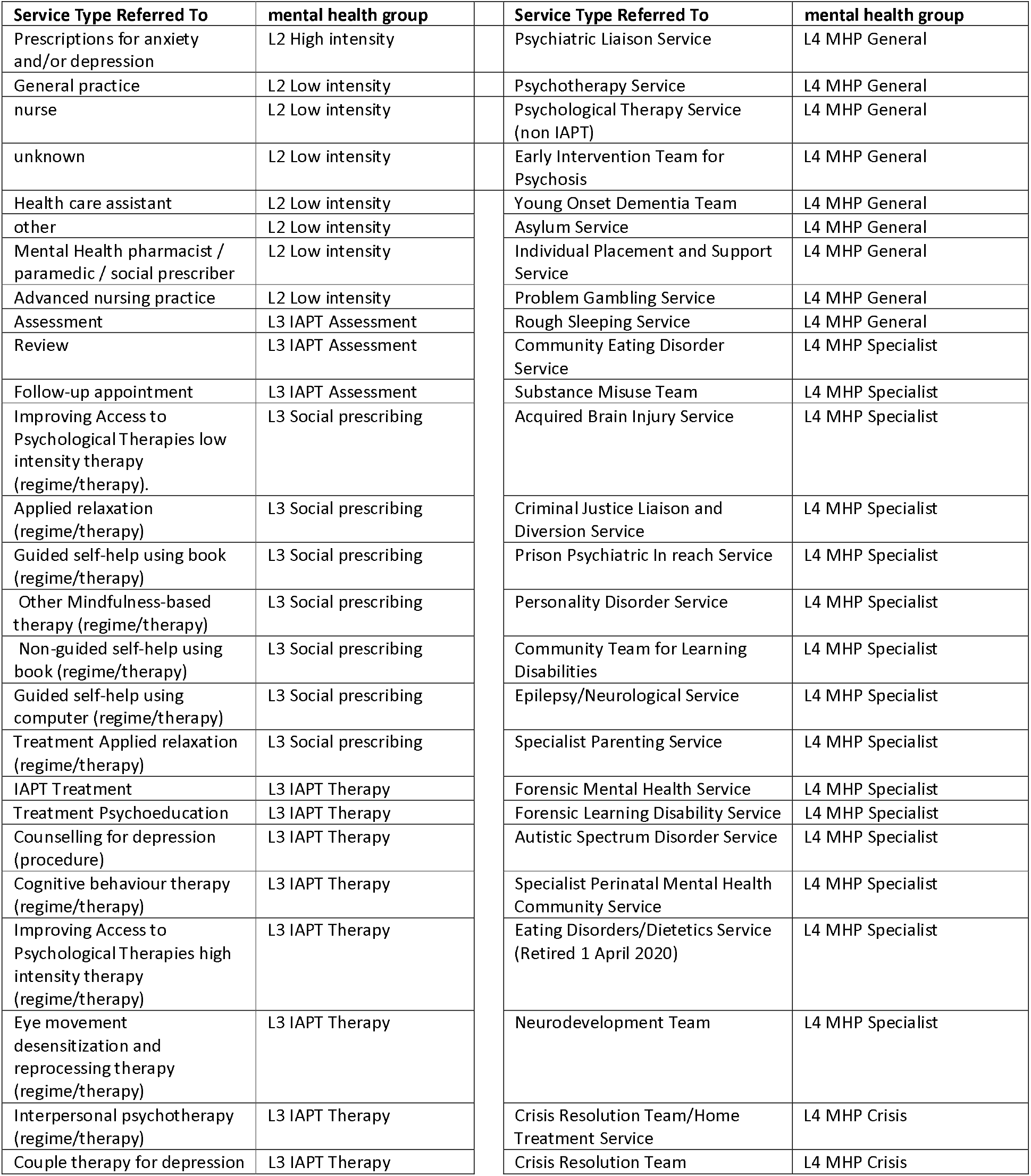
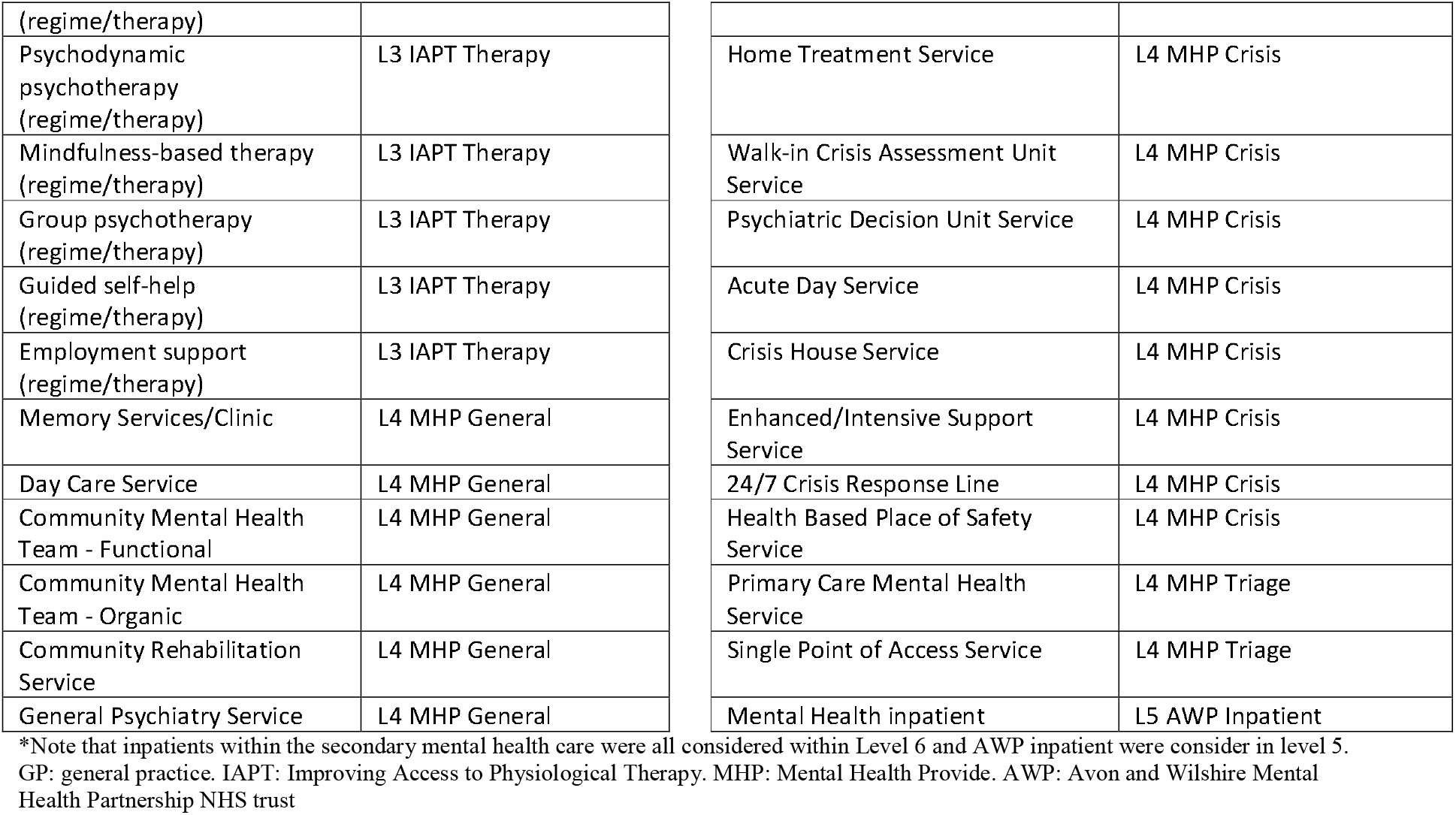
Services type for primary and secondary care mental health outpatients services.

**Figure S1:**
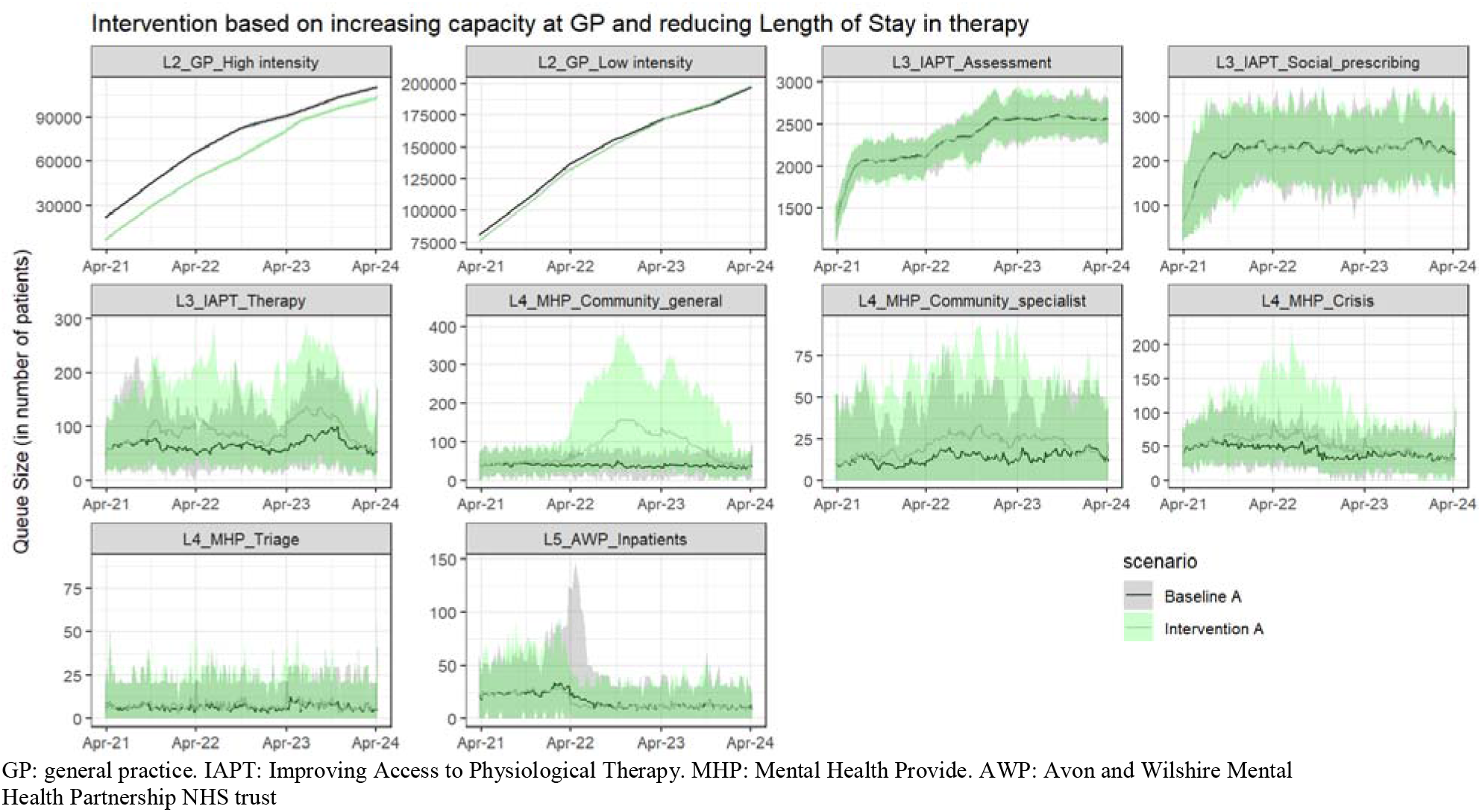
Summary of Weekly Queue size for services modelled post ‘lockdown’ (mean and 95% confidence bands) for the simulation scenarios A.

**Figure S2:**
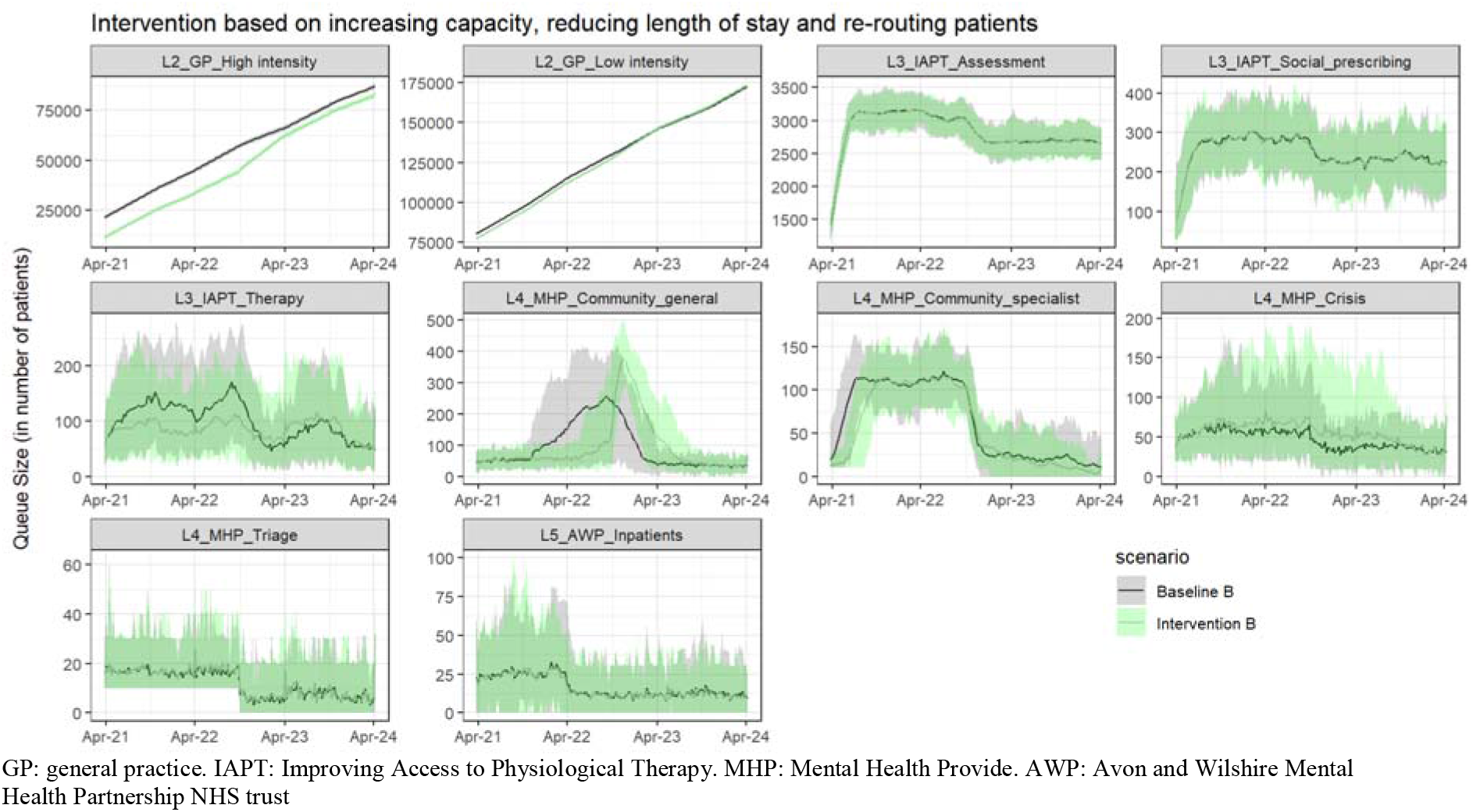
Summary of Weekly Queue size for services modelled post ‘lockdown’ (mean and 95% confidence bands) for the simulation scenarios B.

